# A Canadian Perspective on Family Medicine Residents’ Attitudes and Practices Toward Infants’ Oral Health

**DOI:** 10.64898/2026.01.31.26345294

**Authors:** AlWaleed Abushanan, Quynh Doan, Jolanta Aleksejuniene, Mario Brondani

## Abstract

**Objectives:** To explore the extent to which infant oral health is addressed within the family medicine residencies in Canada, and the attitudes and practices of Canadian family medicine residents towards infant oral health.

**Methods:** Two brief self-administered online surveys, one to 17 Canadian family medicine training program directors and another to current residents within these programs, were conducted using Research Electronic Data Capture (REDCap). Questions focused around respondents’ attitudes and practices towards infants’ oral health and infant oral health content within family medicine curricula.

**Results:** 11 family medicine directors and 155 family medicine residents responded to the survey. 90% of the directors indicated that clinical oral screening was not incorporated in the curriculum. 53% of the residents reported that they did not feel their training was adequate to identify dental caries. 41% described the quality of their oral health training to be poor. While 72% reported lack of knowledge and training as the major barrier to performing oral health-related practices.

**Conclusion:** Most of the family medicine training programs in Canada do not include infant oral health screening in their curriculum. The family medicine residents’ reported lack of knowledge and training is preventing them from performing various oral health-related practices.

## Introduction

Early childhood caries (ECC) is “*the presence of 1 or more decayed (noncavitated or cavitated lesions), missing (due to caries), or filled tooth surfaces in any primary tooth in a preschool-age child (i*.*e. 71 months of age or younger)*” and remains a major oral health problem [1]. ECC is the most common childhood disease; it is 5 times more prevalent than asthma and it often accompanied by serious comorbidities affecting children’s quality of life, their families, the community and the health care system as a whole. It is estimated that 57% of Canadian children 6 to 11 years of age have experienced dental decay, with an average of 2.5 teeth affected by decay per child; in some disadvantaged Indigenous communities the prevalence of decay exceeds 90% [3]. Treatment for ECC under general anesthesia is the most common day surgery at most pediatric hospitals in Canada, it costs annually more than $21 million to the tax payers, and burdens the child’s quality of life [1][4]. Despite its high prevalence, ECC is highly preventable through dietary changes, oral hygiene, and topical fluoride therapies, for example.

The American Academy of Pediatric Dentistry (AAPD) and the Canadian Dental Association (CDA) recommend that dental assessment of infants should be carried out within 6 months of the eruption of the first tooth, no later than year 1 of age [1, 5]. Given that the Canadian dental care delivery system remains mostly privately financed and administered and limiting access to dental care [3], primary care physicians can play a major role in preventing and early detecting ECC [6-9]. In fact, by the age of 3, a child can be seen more than 10 times by a family physician during the well-baby visits assessments for major milestones; by the age of 3, a child might never be seen by a dental professional [6, 10].

In turn, it is essential to convey to primary health care professionals the knowledge regarding infant oral health care so they can be an allied profession in identifying ECC. These professionals can also play an important role in reducing the incidence of ECC and preventing costly surgical intervention for its treatment. The utilization of interdisciplinary collaboration is much needed to further enhance children’s oral health and have a positive influence on their quality of life [11]. Integrating an oral health component in the residency training of primary health care professionals’ curriculum has been recommended [6, 10, 12-14]. In fact, studies from the United States [8, 9, 15-22], Europe [23-27], India [12, 28], Belgium [24], and Turkey [23] have examined the knowledge, attitudes, and practices of primary care physicians towards infant oral health and concluded that, for most part, allied health care providers would be able to identify children with carious teeth, and apply fluoride varnish, for example, after receiving proper training in infant oral health [18]. In Canada, similar investigations are sparse [6]. The objectives of the study were to explore the extent to which infant oral health is addressed within the family medicine residencies in Canada, and the attitudes and practices of Canadian family medicine residents about infant oral health.

## Methods

Two brief self-administered online surveys, one targeting 17 Canadian family medicine training program directors and another targeting the currently enrolled residents within these programs, were conducted suing the Research Electronic Data Capture - REDCap. The directors’ survey consisted of 19 multiple choice, open-ended, agreement/ disagreement, yes/ no questions, while the residents survey consisted of 23 questions of similar design; both surveys were developed from the existing literature and from informal conversations with pediatricians and public health specialists. The content of the surveys included integration of oral health-related subjects in the curriculum, and the residents’ attitudes and practices towards the infants’ the oral health along with some demographic characteristics of the training programs and the residents. A pilot study was conducted among five graduate dental students and five pediatrics residents at the University of British Columbia who evaluated the questions of the survey and suggested adding a question about the number of children seen during rotations and removing three redundant questions. Ethics approval was obtained from the University of British Columbia Research Ethics Board # H16-01582. **A** standardized e-mail to potential participants containing a unique link to the online survey questionnaire was sent via REDCap. Upon clicking the link, the participant received a consent cover letter explaining the nature of the survey along with the questionnaire. Participation was voluntary and no financial incentive for survey completion was offered. Data collection took place between July and September 2018. Although participation status was tracked using unique links, there was no attempt to identify individual participants or to match their responses with the respective program; surveys were coded numerically. The sample size for the residents was estimated at 388, but it was unknown the number of residents who received the email containing the survey link out of the estimated 3217 residents enrolled within the College of Family Physicians of Canada (CFPC) in 2018. All the 17 program’s directors made up the full sample. De-identified demographic information was asked for characterization of the participants only.

Descriptive analysis was computed. Bivariate and multivariate analyses were performed to assess the association between family medicine residents’ attitudes and oral health-related practices and factors such as their postgraduate year, children seen per month, dental caries observed during well-baby visits, quality of training, perceived barriers to carrying out preventive dental practices. Non-parametric analyses (Chi-Square test, and Fischer’s Exact), and binary logistic regression test were performed for categorical variables. The variables of interest for the Directors’ survey were related to the total hours of didactic oral health training the residents receive during their family medicine training, the methods of evaluation of the residents’ oral health knowledge, the perceived barriers towards teaching oral health related topics, the oral health-related topics included in the family medicine curriculum and the oral health-related topics taught in during well-baby visits. The independent variables for the residents’ survey were related to age, gender, the postgraduate year they are in, the frequency of children seen per week, the frequency of which they see children with tooth decay, the quality of the training received, their perceived barriers towards performing oral health-related practices, availability of dental coverage, the availability of shared primary care services with pediatricians, and their attitudes towards (the use of fluoride toothpaste, importance of primary dentition, adequacy of training on oral health, and importance of physicians’ role on promoting oral health). The outcome variables for the residents’ surveys were various oral health-related practices. In the bivariate analyses, the importance of the physicians’ role was dichotomized as not/less important (strongly disagree/ strongly disagree/ neutral) and important (agree, strongly agree). Similarly, for the multivariate analyses, the responses ‘good and excellent’ and ‘disagreed, strongly disagreed and neutral’ were collapsed together due to low number of responses. The statistical analyses were performed using SPSS (IBM Corp. Released 2013. IBM SPSS Statistics for Macintosh, Version 22.0. Armonk, NY: IBM Corp.) and conducted at a confidence interval (CI) of 95%, and a significance level of 0.05.

## Results

A total of eleven program directors completed the survey (64% of response rate). Although the number of residents who actually received the survey is unknown, one hundred and fifty-five responses were received (less than <1% of response rate given the original estimated enrolment number of 3217 residents).

From the directors’ responses, ten (90%) indicated that clinical oral health screening was not incorporated in the curriculum, while 72% (n=8) of them indicated that they curriculum did not include dental disease prevention, early intervention, and ECC. Similarly, the interaction between oral health and general health as well as pregnancy and oral health was not included in 72% of the programs (n=8). All directors indicated that fluoride varnish was discussed in the curriculum. More than half of the directors (n=6) reported that their residents received from 1 to 3 hours of didactic information on oral health-related topics during their entire training. Nearly two thirds (63%, n=7) of the directors indicated that the visual examination of the oral cavity and the child’s teeth and the child’s risk assessment of tooth decay were taught didactically only (Figure 1). Over half the directors (54%, n=6) indicated that they only didactically taught their residents on nocturnal bottle-feeding, on discussing fluoride toothpaste with the parents/caregivers, and on referring children to a dentist. About 63% of them (n=7) indicated that their residents received both didactically and clinically information on the application of topical fluoride products, while 45% of them (n=5) also reported teaching their residents on prescribing fluoride supplements (Figure 1). The most common barriers reported by family medicine directors towards teaching infant oral health-related practices to their residents were competing priorities, knowledge, and family physician’s scope of practice and time (63%, n=7). Furthermore, the majority of directors (81%, n= 9) indicated that they have no method of evaluation of their resident’s oral health knowledge.

Female residents comprised the majority of the respondents (83%, n=128). About 52% (n=80) of the respondents were in their second year; mean age of the participants was 29 years. The vast majority of the residents (90%, n=140) reported that they utilized well-baby visit assessments such as the Rourke Baby Record. The majority of the residents (75%, n=117) seldom saw children younger than 3 years of age, and 76% of them (n=118) had not seen a child with tooth decay. A total of 96 (62%) of the respondents seldom performed a visual examination of the teeth of child between the ages of 1 and 3, while nearly half (48%, n=75) of the residents frequently counseled parents or caregivers regarding teething and dental care; not more than 46% of the respondents (n=72) reported advising the parents or caregivers on teeth cleaning methods (Table 1). Over half the respondents (53%, n=82) assessed the child’s risk for developing tooth decay. When asked about discussing the use of fluoride toothpaste with the parents or caregivers, and referring children to a local dentist, the residents reported that they seldom did these activities (40%, n=62; 43%, n=67, respectively). Around 52% of the residents (n=81) indicated that if dental caries was observed they would have referred the child to the dentist. Half the respondents (n=79) reported recommending bottle weaning between 12 and 24 months of age.

**Table 1:** Family medicine residents’ oral health-related practices.

| **Oral Health-Related Practices of Residents** | **N (%)** |
| --- | --- |
|  | **Total: 155** |
| **Dental assessment of the child’s teeth** |  |
| Never | 18 (11.6) |
| Seldom | 96 (61.8) |
| Frequently | 29 (18.7) |
| Always | 12 (7.7) |
| **Counsel parents/ caregivers regarding teething and dental care** |  |
| Never | 9 (6.0) |
| Seldom | 48 (31.0) |
| Frequently | 75 (48.0) |
| Always | 23 (15.0) |
| **Caries risk assessment of the child’s teeth** |  |
| Never | 37 (24.0) |
| Seldom | 82 (53.0) |
| Frequently | 28 (18.0) |
| Always | 8 (5.0) |
| **Advise parents/ caregivers on teeth cleaning methods** |  |
| Never | 39 (25.0) |
| Seldom | 72 (46.0) |
| Frequently | 36 (23.0) |
| Always | 8 (5.0) |
| **Discuss the use of fluoride toothpaste with parents and/or caregivers** |  |
| Never | 58 (37.0) |
| Seldom | 62 (40.0) |
| Frequently | 30 (19.0) |
| Always | 5 (3.0) |
| **Dental referral** |  |
| Never | 48 (31.0) |
| Seldom | 67 (43.0) |
| Frequently | 33 (21.0) |
| Always | 7 (5.0) |

**Table 2:** Family medicine residents’ attitudes towards oral health-related statements.

| **Oral Health-Related Statements** | **N (%)** |
| --- | --- |
|  | **Total: 155** |
| **Fluoride toothpaste should be used for children under the age of 3** |  |
| Strongly Disagree | 5 (3.0) |
| Disagree | 36 (23.0) |
| Neither agree or disagree | 48 (31.0) |
| Agree | 51 (33.0) |
| Strongly Agree | 15 (10.0) |
| **Baby teeth are important even though they fall out** |  |
| Strongly Disagree |  |
| Disagree | 0 (0.0) |
| Neither agree or disagree | 0 (0.0) |
| Agree | 3 (2.0) |
| Strongly Agree | 76 (49.0) |
|  | 76 (49.0) |
| **I feel my training is adequate for me to identify dental caries in children** |  |
| Strongly Disagree | 32 (21.0) |
| Disagree | 82 (53.0) |
| Neither agree or disagree | 27 (17.0) |
| Agree | 11 (7.0) |
| Strongly Agree | 3 (2.0) |
| **Physicians have an important role in promoting oral health among infants and toddlers** |  |
| Strongly Disagree | 0 (0.0) |
| Disagree | 1 (1.0) |
| Neither agree or disagree | 13 (8.0) |
| Agree | 94 (61.0) |
| Strongly Agree | 47 (30.) |

### Error! Reference source not found

shows the attitudes of family medicine residents towards oral health-related statements. One third of the residents (33%, n=51) *agreed* that fluoride toothpaste should be used for children under the age of 3, while 49% (n=76) *strongly agreed* that primary teeth were important even if they exfoliate. When asked if physicians have an important role in prompting oral health among infants and toddlers, 61% (n=94) of the residents *agreed*. Over half the respondents (53%, n=82) reported that they did not feel their training was adequate to identify dental caries in children (**Error! Reference source not found**.). More than 40% (n=63) of the family medicine residents reported that the quality of their training in oral health-related topics was poor and 39% (n=60) stated that they did not receive any oral health-related training. Furthermore, when asked about potential barriers to perform oral health-related practices, the majority (72%, n=112) of the respondents reported a lack of knowledge or training. Additionally, the vast majority (95%, n=148) of the residents reported that they would like to have more information/resources on identifying dental conditions and disease in infants. They further indicated that they specifically would like to know more about ECC (90%, n=140), and oral home care (86%, n=133).

Bivariate analysis showed significant association between residents who felt that physicians had an important role in promoting oral health to infants and toddlers, and who counseled parents or caregivers regarding teething and dental care (p=0.006), and who discussed fluoride toothpaste use with parents or caregivers (p=0.023) (Table 3). There was a significant association between family medicine residents’ attitudes towards the importance of primary dentition and various oral health-related practices. There was also a significant association between residents who felt that primary dentition was important and who performed an assessment of the child’s teeth (p= <0.001), counseled parents (p=0.008), assessed the child’s risk for developing caries (p=0.001), advised the parents on teeth cleaning methods (p=0.003), and discussed the use of fluoridated toothpastes (p=0.025). There was no significant association between the residents’ attitudes and referrals to a dentist (Table 3). When comparing the residents’ reported frequency of seeing children between 1 and 3 years of age and their oral health-related practices, no significant association was found (data not shown). Table 4 shows a significant association between residents who reported observing dental caries and performing an assessment of the child’s teeth (p=0.003), as well as referring children to a dentist (p=0.047). The reported year of training (PGY1, PGY2) indicated that residents in their first year were less likely to visually examine the child’s teeth (p=0.035) and discuss the use of fluoridated toothpastes with the parents (p=0.050) than those in the second year of their training (Table 4). There were also no significant differences in any of the practice patterns between the residents perceiving different barriers for their oral health-related practices (data not shown).

**Table 3:** Association between physicians with different attitudes about the importance of physicians’ role in promoting oral health and primary dentition and oral health-related practices.

| PRACTICE PATTERNS | PHYSICIAN'S ROLE |  | Significance<sup>#</sup> |
| --- | --- | --- | --- |
|  | NO<br>N (% of total) | YES<br>N (% of total) |  |
| **Dental assessment the child's teeth** |  |  |  |
| Never/seldom | 12 (85.7) | 102 (72.3) | 0.279 |
| Frequently/always | 2 (14.3) | 39 (27.7) |  |
| **Parental counseling** |  |  |  |
| Never/seldom | 10 (71.4) | 47 (33.3) | **0.006\*** |
| Frequently/always | 4 (28.6) | 94 (66.7) |  |
| **Caries risk assessment** |  |  |  |
| Never/seldom | 12 (85.7) | 107 (75.9) | 0.406 |
| Frequently/always | 2 (14.3) | 34 (24.1) |  |
| **Advise parents about tooth cleaning** |  |  |  |
| Never/seldom | 13 (92.9) | 98 (69.5) | 0.065 |
| Frequently/always | 1 (7.1) | 43 (30.5) |  |
| **Discuss the use of fluoride** |  |  |  |
| Never/seldom | 14 (100.0) | 106 (75.2) | **0.023\*** |
| Frequently/always | 0 (0.0) | 35 (24.8) |  |
| **Dental referral** |  |  |  |
| Never/seldom | 13 (92.9) | 102 (72.3) | 0.094 |
| Frequently/always | 1 (7.1) | 39 (27.7) |  |
| PRACTICE PATTERNS | IMPORTANCE OF PRIMARY DENTITION |  | Significance<sup>#</sup> |
|  | PGY1<br>N (% of total) | PGY2<br>N (% of total) |  |
| **Dental assessment the child's teeth** |  |  |  |
| Never/seldom | 69 (87.3) | 45 (59.2) | **<0.001\*** |
| Frequently/always | 10 (12.7) | 31 (40.8) |  |
| **Parental counseling** |  |  |  |
| Never/seldom | 37 (46.8) | 20 (26.3) | **0.008\*** |
| Frequently/always | 42 (53.2) | 56 (73.7) |  |
| **Caries risk assessment** |  |  |  |
| Never/seldom | 69 (87.3) | 50 (65.8) | **0.001\*** |
| Frequently/always | 10 (12.7) | 27 (34.2) |  |
| **Advise parents about tooth cleaning** |  |  |  |
| Never/seldom | 65 (82.3) | 46 (60.5) | **0.003\*** |
| Frequently/always | 14 (17.7) | 30 (39.5) |  |
| **Discuss the use of fluoride** |  |  |  |
| Never/seldom | 67 (84.8) | 53 (69.7) | **0.025\*** |
| Frequently/always | 12 (15.2) | 23 (30.3) |  |
| **Dental referral** |  |  |  |
| Never/seldom | 61 (77.2) | 54 (71.1) | 0.381 |
| Frequently/always | 18 (22.8) | 22 (28.9) |  |

**Table 4:**
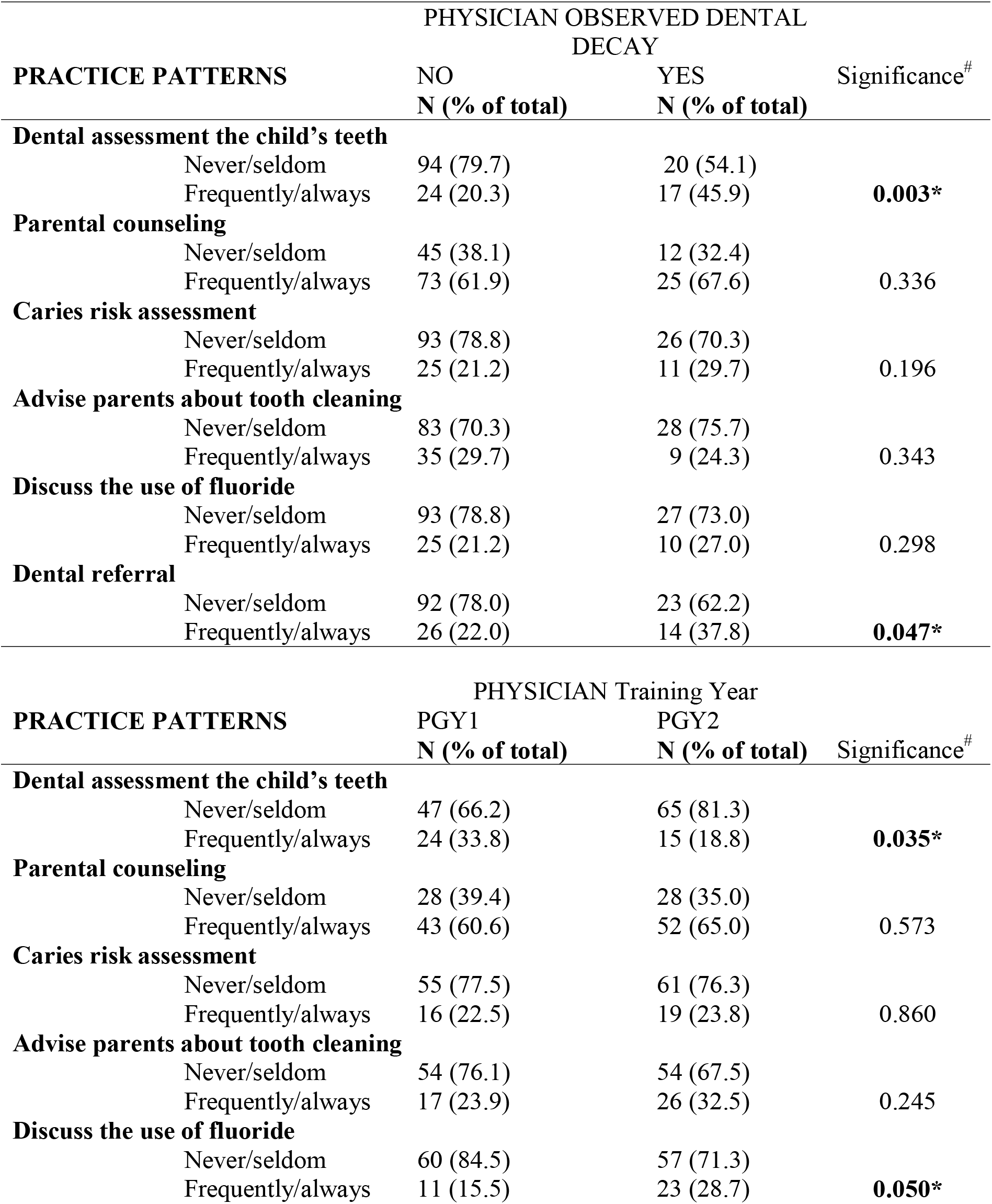
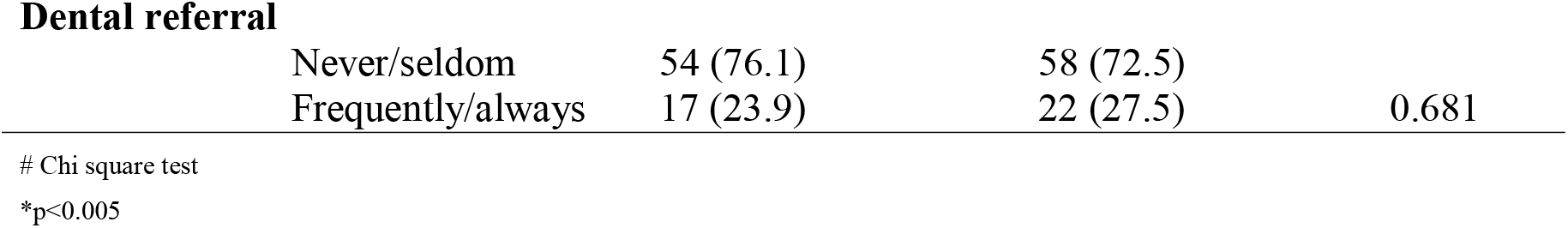
Association between physicians who observed and did not observe caries, year of training and oral health-related practices.

| PRACTICE PATTERNS | PHYSICIAN OBSERVED DENTAL<br>DECAY |  | Significance<sup>#</sup> |
| --- | --- | --- | --- |
|  | NO<br>N (% of total) | YES<br>N (% of total) |  |
| **Dental assessment the child's teeth** |  |  |  |
| Never/seldom | 94 (79.7) | 20 (54.1) | **0.003\*** |
| Frequently/always | 24 (20.3) | 17 (45.9) |  |
| **Parental counseling** |  |  |  |
| Never/seldom | 45 (38.1) | 12 (32.4) | 0.336 |
| Frequently/always | 73 (61.9) | 25 (67.6) |  |
| **Caries risk assessment** |  |  |  |
| Never/seldom | 93 (78.8) | 26 (70.3) | 0.196 |
| Frequently/always | 25 (21.2) | 11 (29.7) |  |
| **Advise parents about tooth cleaning** |  |  |  |
| Never/seldom | 83 (70.3) | 28 (75.7) | 0.343 |
| Frequently/always | 35 (29.7) | 9 (24.3) |  |
| **Discuss the use of fluoride** |  |  |  |
| Never/seldom | 93 (78.8) | 27 (73.0) | 0.298 |
| Frequently/always | 25 (21.2) | 10 (27.0) |  |
| **Dental referral** |  |  |  |
| Never/seldom | 92 (78.0) | 23 (62.2) | **0.047\*** |
| Frequently/always | 26 (22.0) | 14 (37.8) |  |

| PRACTICE PATTERNS | PHYSICIAN Training Year |  | Significance<sup>#</sup> |
| --- | --- | --- | --- |
|  | PGY1<br>N (% of total) | PGY2<br>N (% of total) |  |
| **Dental assessment the child's teeth** |  |  |  |
| Never/seldom | 47 (66.2) | 65 (81.3) | **0.035\*** |
| Frequently/always | 24 (33.8) | 15 (18.8) |  |
| **Parental counseling** |  |  |  |
| Never/seldom | 28 (39.4) | 28 (35.0) | 0.573 |
| Frequently/always | 43 (60.6) | 52 (65.0) |  |
| **Caries risk assessment** |  |  |  |
| Never/seldom | 55 (77.5) | 61 (76.3) | 0.860 |
| Frequently/always | 16 (22.5) | 19 (23.8) |  |
| **Advise parents about tooth cleaning** |  |  |  |
| Never/seldom | 54 (76.1) | 54 (67.5) | 0.245 |
| Frequently/always | 17 (23.9) | 26 (32.5) |  |
| **Discuss the use of fluoride** |  |  |  |
| Never/seldom | 60 (84.5) | 57 (71.3) | **0.050\*** |
| Frequently/always | 11 (15.5) | 23 (28.7) |  |
| **Dental referral** |  |  |  |
| Never/seldom | 54 (76.1) | 58 (72.5) | 0.681 |
| Frequently/always | 17 (23.9) | 22 (27.5) |  |

Table 5 presents the analysis of the explanatory variables associated with the residents performing dental assessments on children and counselling the parents or caregivers. This logistic regression model indicates that the residents who reported their training quality as poor were significantly less likely to perform a dental assessment of the children’s teeth (OR=0.38; CI=0.15, 0.99). Furthermore, residents who strongly agreed to the importance of their role in promoting oral health to infants were 6 times more likely to perform a dental assessment (OR=6.09; CI=1.12, 33.0). Residents who strongly agreed to the importance of promoting oral health to infants were significantly 4 times more likely to provide oral health counselling (OR=4.91; CI=1.21, 19.8).

**Table 5:**
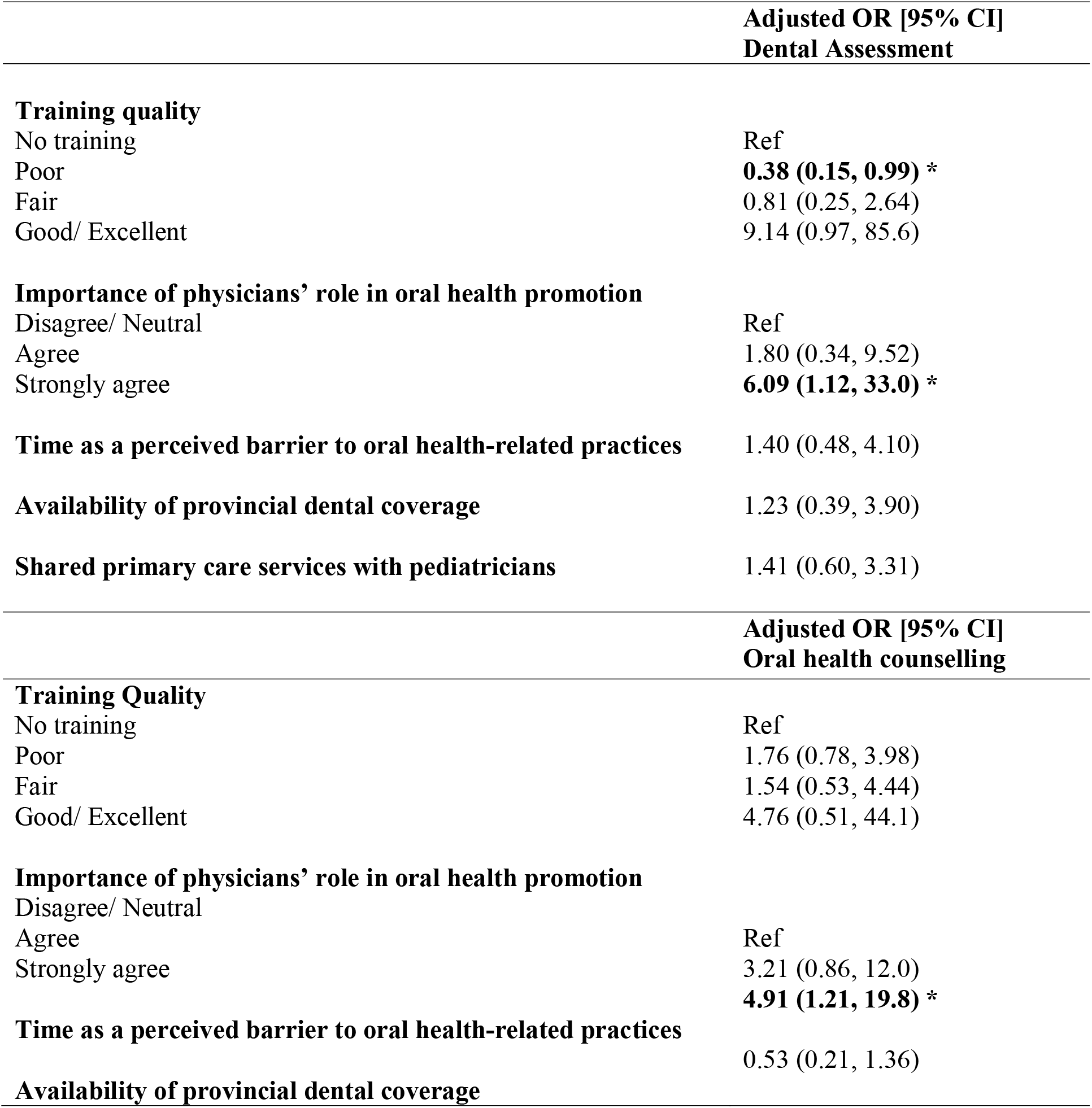
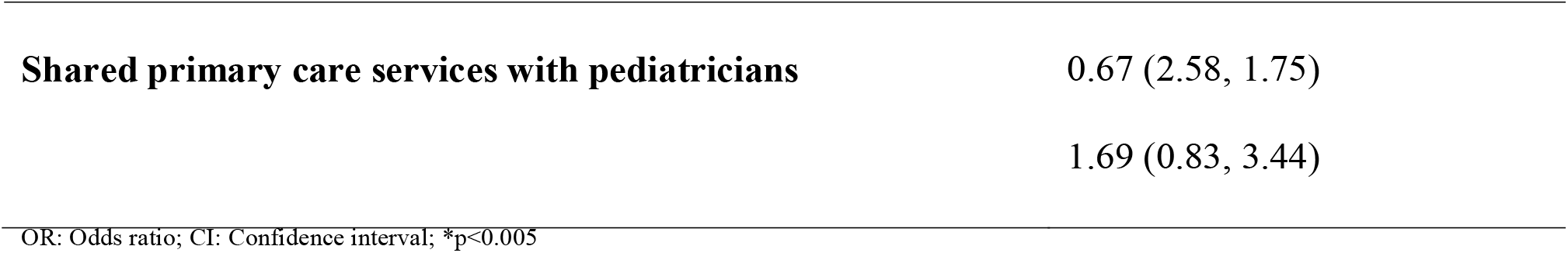
Multivariate logistic regression model of variables associated with dental assessment and oral health counselling.

## Discussion

The results of this study indicated that there is a lack of oral health education within the family medicine residents’ training in Canada, which is in concurrence with the existing literature. The majority of the directors stated that their curriculum included didactic sessions on how to visually examine children’s teeth, assess their caries risk, nocturnal bottle feeding, and discuss the use of fluoridated toothpastes with the child’s caregiver. On the other hand, the 1 to 3 hours allocated to infant oral health may not be enough to cover more of the clinical oral health screening, dental disease prevention and intervention, ECC, interaction between oral and general health, pregnancy and oral health and fluoride varnish. A previous study reported that oral health education in a medical setting need to be designed in an efficient way, as didactic teaching alone does not suffice to ensure efficacy of the oral health training [25]. Most of the directors reported competing priorities, lack of knowledge and time as their perceived barriers to teaching these oral health-related topics. A recent study by Silk et al. reported similar findings as only half the programs reported having up to 3 hours of oral health component [29]. They also reported similar barriers to having an oral health curriculum as reported in this study. Similarly, Prakash et al. investigated practicing Canadian family physicians in 2006, and found that one third of them reported the same 1 to 3 hours of oral health training; the majority of the programs had either no oral health training or felt that it was poor [6]. This finding can be an indication that in the past 12 years there has been no change to the oral health training in family medicine residency programs in Canada. The vast majority of the directors reported no methods of evaluation of their residents’ oral health knowledge. The residents’ responses can be reflective of the aforementioned findings from the directors, as over half the residents reported a feeling of inadequacy towards their training in identifying dental caries. Furthermore, the findings reported herein were supported by the perceived need for more information/resources in dental diseases (95%, n=148), with the most respondents identifying ECC and oral home care as two topics they would like to know more about as also found be [6]. The lack of oral health training makes it difficult for family medicine residents and physicians to help curbing the impact of untreated ECC upon their infant patients and their families. The various competing priorities, and lack of faculty expertise in oral health makes it challenging to have a positive oral health involvement in the residents’ training. Although 90% of the respondents use well-baby visits assessment such as the Rourke Baby Record, it remains unknown the extent to which the ‘*other’* section containing oral health information is actually used. Moreover, many residents reported the lack of resources or guidelines as perceived barriers to be comfortably perform many of the oral health-related practices.

Almost 40% of the residents agreed with the importance of fluoridated toothpastes in children younger than 3 years, while the other 60% did not agree. Such discrepancy might be explained by conflicting resources currently available. For instance, the Canadian Pediatric Association’s Caring for Kids website recommends the use of fluoridated toothpastes only after 3 years of age [30], while the CDA recommends its use in children younger than 3 years based on the caries risk [31].

The findings also showed that the residents who felt that they hold an important role in promoting oral health amongst infants and toddlers were also more likely to counsel parents on teething and dental care, to discuss the use of fluoride, and to recognize the importance of primary dentition even though they exfoliate. These findings were in concurrence with previous studies in the literature [6, 32]. Although it is reassuring that the vast majority of family medicine residents’ attitudes were positive towards various oral health-related topics, their lack of training and knowledge might be a hindering factor in fully integrating oral health prevention and promotion into their practices.

The majority of residents did not perform oral health-related practices during well-baby visits; very few residents saw children 3 years or younger. This can be explained by the fact that family medicine residents might have different clinical rotations throughout their residency and depending on their year of training, they might not see children at all. Furthermore, depending on the province of which the residents are training, pediatricians might be seeing more children than family physicians [30]. Over two thirds of the residents reported never observing dental caries on children 0-3 years of age, which can be explained by the fact that the majority of the respondents did not see children and when they saw, they might not have lifted the lip or know what they were looking for due to lack of knowledge and training. However, residents also indicated that if dental caries was observed, they would have advised the parents/caregivers to take the child to the dentist. Family physicians remain a valuable allied workforce to accurately assess and refer children with ECC for proper care [18], but more importantly, in helping to prevent ECC [22]. There are several limitations to this study. The residents’ response rate was low (n=155) given that it was difficult in accessing all family medicine residents across Canada as the CFPC does not endorse third-party or external project surveys to its members. Also, the survey did not have a French version which limited our accessibility to French speaking universities and residents. In terms of the limitations with the survey itself, there was the potential for residents completing the survey based on what they believed the investigators would like to know or what they believed was right to answer. Although attempts were made to ensure confidentiality and anonymity of the respondents, the generalizability of our study remains limited. The survey was based on self-reported answers; therefore, this might not be an accurate representation of the actual training received and the practices performed by the residents.

For the directors’ survey, the 11 responses limit generalizability to all residency programs in Canada or elsewhere. Future studies in collaboration with the CFPC are recommended to better understand the gaps in oral health training and education in family medicine residencies. On the other hand, studies should also include the extent to which general dental professionals, not dental pediatricians, are comfortable in seeing infants and children younger than 6 years-old once they are referred from family physicians.

## Conclusions

It is essential to convey to primary health care professionals the knowledge regarding infant oral health care so they can be an allied profession in identifying ECC. However, most of the family physicians training programs in Canada do not include infant oral health screening in their curriculum comprehensively. While the majority of family medicine residents felt that physicians have an important role in promoting oral health amongst children, the reported lack of knowledge and training is hindering them from performing various oral health-related practices. Such lack of knowledge can be attributed to their perceived lack of training in oral health. Efforts should be made to increase interprofessional collaboration between dentistry and family medicine curricula; this in turn will help reduce the incidence and severity of ECC.

## Data Availability

All data produced in the present study are available upon reasonable request to the authors.

## Notes

### Competing Interest Statement

The authors have declared no competing interest.

### Funding Statement

This study did not receive any funding

### Author Declarations

Ethics approval was obtained from the University of British Columbia Research Ethics Board H16-01582.

